# Threat imminence reveals links among unfolding of anticipatory physiological response, cortical-subcortical intrinsic functional connectivity, and anxiety

**DOI:** 10.1101/2021.08.21.21262409

**Authors:** Rany Abend, Sonia G. Ruiz, Mira A. Bajaj, Anita Harrewijn, Julia O. Linke, Lauren Y. Atlas, Daniel S. Pine

## Abstract

Excessive expression of threat-anticipatory defensive responses is central in anxiety. Animal research indicates that anticipatory responses are dynamically organized by threat imminence and rely on conserved circuitry. Insight from translational work on threat imminence could guide mechanistic research mapping abnormal function in this circuitry to aberrant defensive responses in anxiety. Here, we initiate such research.

Fifty pediatric anxiety patients and healthy-comparisons (33 females) completed a threat-anticipation task whereby cues signaled delivery of highly-painful (threat) or non-painful (safety) heat. Temporal changes in skin-conductance indexed defensive responding as function of threat imminence. Resting-state functional connectivity data were used to identify intrinsic-function correlates of anticipatory response within a specific functional network derived from translational research.

Results indicate that anxiety was associated with greater increase in anticipatory response as threats became more imminent. Magnitude of increase in threat-anticipatory responses corresponded to intrinsic connectivity within a cortical-subcortical circuit; importantly, more severe anxiety was associated with greater connectivity between ventromedial prefrontal cortex and hippocampus and basolateral amygdala, a circuit implicated in animal models of anxiety. These findings link basic-translational and clinical research, highlighting aberrant intrinsic function in conserved defensive circuitry as potential pathophysiological mechanism in anxiety.

## Introduction

Anticipating an encounter with threat elicits a cascade of defensive behaviors that unfold with *imminence*, or proximity, of the threat. While threat-anticipatory defensive responses clearly possess an adaptive value, their excessive expression is a hallmark characteristic of pathological anxiety^1-5^. The conserved nature of defensive responses can guide research on their aberrant expression in anxiety. To date, such research has not considered threat imminence to delineate aberrant dynamic threat-anticipation processes in anxiety and its underlying pathophysiological neural mechanisms. In this report, we examine whether anxiety is associated with distinct patterns of defensive responding as threat becomes increasingly imminent, and link these response patterns to function in conserved neural circuitry.

Extensive research in animals indicates that defensive behaviors are dynamically organized by *threat imminence* and available behavioral options^1,2,6-9^. For example, when a threat appears (a phase referred to as threat *encounter*/*post-encounter*), animals typically display freezing or passive avoidance behavior to evade detection and assess risks; as the threat looms closer and attack is imminently anticipated (*circa-strike* phase), acute defensive behaviors, such as active avoidance, become more prominent^2,5,8,10,11^. Distinct patterns of physiological responding, such as changes in heart rate and skin conductance, accompany such defensive behaviors, promoting their execution^3,5,12,13^. Given the adaptive value of threat-anticipatory defensive responses and their conserved underlying brain circuitry, it has been proposed that such responses in humans are likewise determined by threat imminence. Indeed, recent work suggests that increasing threat imminence is associated with shifts to more acute defensive behaviors and physiological responses^3,8,14-19^.

Excessive threat-anticipatory behavioral, physiological, and cognitive responses are central in the presentation of anxiety symptoms^20,21^. Thus, from an evolutionary-translational perspective, pathological anxiety could reflect aberrant function in the circuitry generating threat-anticipatory defensive responses, leading to their maladaptive expression^5,8,22,23^. Important insight has been provided by studies that index average or maximal threat-anticipatory physiological responses in conditioning/extinction learning procedures or contexts of unpredictable delivery of aversive stimuli^5,22,24^. However, considering the imminence of the threat, and corresponding defensive responses, may importantly clarify the temporal dynamics and organization of anxiety symptoms. For example, distal threats may elicit excessive cognitive responses, such as pathological worry and inaccurate risk assessment; as threat becomes imminent, anxiety manifests as excessive reactive, acute responses associated with panic states, such as acute changes in physiological response and increased active avoidance behavior^1,8,21,23,25^. Research guided by insight from animal work on characterizing the dynamic nature of defensive responding could therefore inform more precisely on the dynamics of psychopathological defensive processes expressed as anxiety symptoms. Our first aim examines links between anxiety severity and patterns of defensive responding as a function of threat imminence.

Research in animals consistently links the selection and execution of defensive responses to function within a specific, distributed network of brain structures that includes amygdala nuclei, hippocampus, hypothalamus, and periaqueductal gray (PAG), and infralimbic and prelimbic cortices^1,2,8,9,19,26^, referred to henceforth as “defensive response” circuitry. Recent work begins to translate such findings on threat imminence and this circuitry to human-subject paradigms^3,8,14-19,27^, but precise characterization of the functional links within it is still limited. Given emerging cross-species findings, we hypothesize that excessive defensive responding observed in anxiety relates to interindividual differences in the functional organization of this circuit, and particularly cortical-subcortical connections driving the selection, maintenance, and regulation of imminence-specific defensive responses^8,22,28-30^. Linking maladaptive patterns of conserved defensive responses to pathological anxiety could promote translational neuroscience-guided research on pathophysiological mechanisms and potential biomarkers.

From a biomarker standpoint, there is considerable value in identifying individual differences in the *intrinsic* function of the defensive response network that correlate with its aberrant expression in anxiety; such associations could potentially help identify individuals prone to express excessive defensive responses across contexts, a key characteristic of anxiety^20,21^. Resting-state functional connectivity (rsFC), reflecting correlated spontaneous neural activity fluctuations across functionally-related regions, could be used to identify such intrinsic variations in network function. rsFC data are increasingly used to characterize the intrinsic organization of distributed networks^31-33^ with high reliability and sensitivity to inter-subject differences^34-36^. Growing evidence suggests that psychopathology manifests in aberrant rsFC patterns, potentially indicative of intrinsic perturbations or biases in specific functional networks^37-44^. Such biases may thus serve as potential markers and treatment targets for pathological neurobiological processes^44-49^. Our second aim explores links between imminence-dependent defensive responding in anxiety and variations in rsFC-derived intrinsic function within the defensive response network.

In this report, we induced threat anticipation in healthy and clinically-anxious youth by cues instructed to signal the subsequent delivery of highly painful (threat) or non-painful (safety) thermal stimulation. We assessed the temporal unfolding of anticipatory defensive responding as a function of threat imminence, indexed by changes in skin conductance levels^3,16,18,50^. Drawing from animal research, we focused on two potential specific anticipatory effects^10,11^: the initial physiological response to cue onset (corresponding to the threat *encounter* phase), and expected increase in physiological response with increasing threat imminence (*circa-strike*). Network analyses on rsFC data from these subjects were then used to link individual differences in defensive responding to intrinsic functional organization of the defensive response circuitry.

Given that anxiety features greater disposition for defensive responses^20^, we hypothesized that more severe anxiety will be associated with stronger initial physiological response and a greater increase in response as threat becomes increasingly imminent. Second, we hypothesized that connectivity within the defensive response circuitry, and specifically in cortical-subcortical connections, will correlate with magnitude of threat-anticipatory responding in both phases. Third, we hypothesized that more severe anxiety will be associated with aberrant cortical-subcortical function within this circuitry as it relates to threat-anticipatory responding, reflecting intrinsic tendency for abnormal selection and regulation of responses to threat^29,30,51,52^.

## Methods

### Participants

Data were from a sample of 50 youth recruited to participate in research on fear and anxiety at the National Institute of Mental Health (NIMH) and which appeared in our previous report^53^. This sample included 25 medication-free, treatment-seeking youth with pediatric anxiety disorders (17 females; *M*age=14.06 years, range=9.34–17.90) and 25 healthy comparisons (HC; 16 females; *M*age=14.90 years, range=9.30–17.28). See supplement for inclusion/exclusion criteria. Of note, we studied youth since anxiety typically emerges in early age and precedes additional, compounding psychopathology^54-56^; studying youth with anxiety and no other disorders enables us to link more tightly observed effects specifically to anxiety.

Groups did not differ in sex, mean age, or mean IQ, *p*s>0.17; the anxiety group reported higher mean current anxiety symptoms (see below), *t*(30.44)=11.87, *p*<0.001. Written informed consent was obtained from parents of participants; written assent was obtained from youth. Procedures were approved by the NIMH Institutional Review Board. Participants received monetary compensation. Patients completed the study prior to treatment. Raw physiological data have been used in our previous report, in which we noted greater averaged anticipatory physiological responding in individuals with anxiety disorders^53^. Importantly, in our prior analyses, we did not consider the chronometry of physiological responses; as such, all physiology indices and analyses reported here, as well as imaging data, are novel.

### Anxiety severity

All participants were carefully assessed by trained clinicians using a structured clinical interview^57^. The anxiety group was composed of treatment-seeking, medication-free youth who received a diagnosis of anxiety disorder and no other psychiatric disorders; the HC group was composed of youth without any psychiatric disorders. Analyses also considered anxiety severity dimensionally across the sample using the parent-reported Screen for Child Anxiety Related Emotional Disorders (SCARED)^58^, to examine dimensional mechanistic variations. This “gold standard” measure of anxiety symptom severity includes 41 items pertaining to anxiety-related symptoms or behaviors; each item is rated on a 3-point Likert-type scale (0=not true, 2=very true); the total score was used in analyses. Mean (SD) total score was 6.39 (3.21) in the HC group and 28.48 (10.68) in the anxiety group.

### Threat-anticipation task

Participants completed a threat anticipation task described in detail in previous work^53,59,60^; see Fig. 1A. Briefly, shape cues were paired with individually-calibrated thermal stimulation applied to the forearm. Participants were instructed that one shape predicted non-painful stimulation (safety) while the other predicted highly painful stimulation (threat). We chose to use noxious thermal stimulation as it is a primary, robust reinforcer signaling potential physical damage. In each trial, participants were presented with the safety or threat cue (2s). Eight seconds after cue onset, participants received the relevant painful or non-painful thermal stimulation (4s, plus 0.5s ramp up and down). After variable delay (5-7s), participants provided pain ratings using a mouse (0-10 scale). Variable inter-trial interval (4-6s) separated trials. Eighteen trials featuring each of the two cues, in counterbalanced order, were administered. See supplement for additional details and data collected in the original task.

**Fig. 1.**
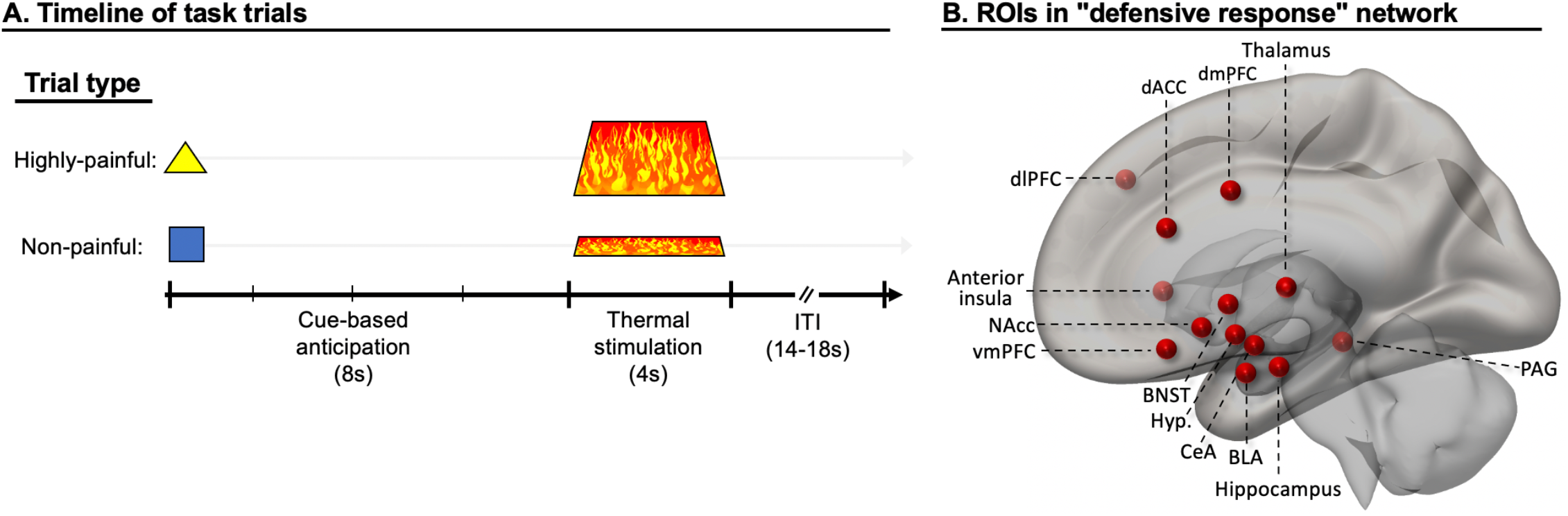
Threat-anticipation task and defensive response network ROIs. A) Timeline of task trials. Each trial started with presentation of cue instructed to predict highly-painful or non-painful thermal stimulation (temperatures individually calibrated before task), initiating an anticipation phase (8s). Next, thermal stimulation was applied (4s), followed by an intertrial interval (ITI; 14-18s) which included pain ratings. B) Literature-guided network of ROIs (regions of interest; here, showing just one hemisphere, in a mid-sagittal view with transparent cortex) selected for targeted analyses linking intrinsic functional connectivity and threat-anticipatory physiological responding. *Note*: dlPFC=dorsolateral prefrontal cortex, dACC=dorsal anterior cingulate cortex, vmPFC=ventromedial prefrontal cortex, dmPFC=dorsomedial prefrontal cortex, BNST=bed nucleus of the stria terminalis, Hyp.=hypothalamus, CeA=central nucleus of the amygdala, BLA=basolateral amygdala, NAcc=nucleus accumbens, hippo=hippocampus, PAG=periaqueductal gray.

### Psychophysiological recording and processing

Skin conductance level (SCL) data were used to assess physiological responding^3,50^, recorded continuously at 1000Hz from the left middle and index fingers using Biopac equipment and AcqKnowledge software (Goleta, CA). Following prior work^53,61^, continuous deconvolution (*Ledalab* MATLAB package) was used to decompose SCL data into tonic and phasic components^62^.

### Data analysis

The combination of instructed contingencies and fixed anticipation window enabled assessment of precise temporal effects in the unfolding of anticipatory defensive responding. Accordingly, analyses focused on changes in physiological response within the 8-second, cue-based anticipation phase starting with cue onset and ending with thermal stimulation delivery. Outcome imminence was operationalized as time between cue appearance and outcome delivery. Within this window, SCL data were assessed in four 2-second, non-overlapping, successive bins. Within each bin we then derived the average SCL signal and subtracted from it the mean tonic SCL level; this served as the index of physiological response in analyses. This measure enabled us to assess the magnitude of physiological responding in each bin while controlling for individual differences in local tonic signal and responsivity, and without making assumptions on response functions or relying on pre-cue epochs (which, by definition, include a pre-encounter anticipation period) to derive a baseline. Tonic response magnitude did not change by bin, *p*=0.50; further, groups did not differ in mean tonic response magnitude, *p*=0.51. Each bin response was then square-root transformed^63^; following transformation, these values did not deviate from the normal distribution (*p*s>0.16, Kolmogorov-Smirnov test). The four bins then comprised the factor of outcome imminence.

To facilitate interpretation, we first considered anxiety as a categorical variable; we then extend analyses to test dimensional effects of anxiety severity across the sample. First, we examined whether anxiety was associated with distinct temporal patterns of threat-anticipatory response with a repeated-measures ANOVA testing the effect of Cue×Imminence×Group on physiological response, with Cue (safety, threat) and Imminence (bins 1-4) as within-subject factors, and Group (HC, anxiety) as a between-subject factor. In line with our hypotheses, we then examined anxiety effects on two specific imminence-derived anticipatory physiological response measures: 1) initial response to threat (bin 1), and 2) magnitude of threat-specific *increase* in physiological response across anticipation window (quantified as response in bin 4 minus response in bin 1, in threat relative to safety trials^1^). These indices were then used in the rsFC analyses (see below).

Significant interactions were decomposed by lower-order tests. Correlations reflect Pearson correlation coefficients. All tests were two-tailed; significance was set at *p*<0.05. Cohen’s *d* (*t*-tests) and partial eta squared (ANOVA) were used to calculate effect sizes.

### Intrinsic connectivity correlates of anticipatory physiological response

Work in animals consistently links threat-anticipatory defensive responses to a specific brain circuitry^1,6,8,52,64-67^. Key elements in this defensive response circuitry include subcortical structures such as basolateral amygdala (BLA), central nucleus of the amygdala (CeA), ventral (anterior in humans) hippocampus, bed nucleus of stria terminalis (BNST), nucleus accumbens (NAcc), midline thalamic nuclei (including the paraventricular nucleus^68^), hypothalamus, and periaqueductal gray (PAG), as well as cortical regions, primarily infralimbic and prelimbic cortices (comparable to vmPFC and dorsal anterior cingulate/dorsomedial prefrontal cortex, respectively), and anterior insula. Research in humans is beginning to extend such findings, indicating the involvement of similar regions, as well as dorsolateral prefrontal cortex (dlPFC), in threat anticipation states^6,14,22,23,26,27,69-72^. However, direct linking of function in this circuitry to physiological and behavioral indices of defensive responding in humans, as is done in animals, remains limited^18,19,27,70,73,74^. Importantly, research linking perturbed function in this circuitry to aberrant defensive responding in anxiety is needed in order to establish its pathophysiological role. Here, we begin to bridge this gap by identifying intrinsic functional connectivity patterns that covary with individual differences in magnitude of threat-anticipatory physiological responses, and the moderation of such associations by anxiety.

### Imaging acquisition and preprocessing

Resting state imaging data (10-minute, multi-echo sequence; see supplement), including field map distortion correction scans, were acquired on a 3T MR750 General Electric scanner (Waukesha, Wisconsin, USA) at the NIMH (*med*=27 days between task and imaging visits). Imaging data from 13 participants of the full sample were not available due to MRI contraindications or unavailability/refusal to scan (5 patients and 8 healthy controls). Functional imaging data were preprocessed and normalized to MNI space with FMRIPrep^75^; see supplement. Data were then entered into CONN software^76^, where they were resampled into 2mm resolution and underwent denoising which included removal (by means of regression) of pre-steady-state outliers, cosine, and all ICA-AROMA time-series regressors^77^, which is particularly suited for network identifiability^78^. Further, standard bandpass filter [0.008-0.09Hz] and linear detrending were applied. Quality control–functional connectivity (QC–FC) correlations for all motion variables indicated 90.7-98.6% overlap with null-hypothesis distribution, indicating effective motion denoising^78^. As suggested^79-82^, unsmoothed data were used in analyses to avoid signal “spillage” between nearby regions of interest (ROIs) which could artificially affect connectivity measures. One participant was excluded from analyses due to excessive motion, leaving a total sample of 36 participants; see supplement for more details.

### Data analysis

We aimed to identify patterns of intrinsic functional connectivity within the defensive response circuitry^82^ that relate to the magnitude of expressed threat-anticipatory physiological response, indexing preparation for execution of defensive responses^2,3^. We focused on two potential effects: the magnitude of *initial* response to threat cues (first bin) and the magnitude of *increase* in anticipatory response across the anticipation window (increase in response from first to last bin).

Guided by the literature^1,2,4,6,26,64,71^, the subcortical ROIs in the defensive response network (see Fig. 1B) included bilateral BLA, CeA, anterior hippocampus, BNST, NAcc, midline thalamus, lateral hypothalamus, and PAG (single ROI). In the cortex, we included ROIs in bilateral vmPFC, dorsal anterior cingulate (dACC), dorsomedial PFC (dmPFC), dlPFC, and anterior insula. All 25 ROIs in this network were defined using in-house scripts that merge several publicly-available, expert cortical and subcortical segmentations into the MNI standard space (https://github.com/rany-abend/atlas); see supplement for more information on ROI selection and definition.

Once this network of 25 nodes was defined, an adjacency matrix of zero-order connectivity correlations between the nodes (300 edges) was computed at the individual subject level. Threat-anticipatory physiological response indices, anxiety severity (assessed continuously using SCARED scores, to increase power), and age (in years) were mean-centered and used as between-subject, second-level covariates. Network-Based Statistics (NBS) second-level analyses were then carried out, as implemented in CONN^76,83^, to pursue our second hypothesis. Thus, to identify specific subnetworks in which connectivity was associated with the magnitude of threat-anticipatory response, analyses tested the correlation between *network mass* (weighted sum of edges) and physiological response magnitude^83^. We used an edge-wise initial uncorrected correlation threshold of *p*<0.005 in conjunction with a threshold of *p_FDR_*<0.05 for network mass; the test statistic was tested relative to 5,000 permutations under the null hypothesis^83^. Finally, to test our third hypothesis, we examined whether anxiety severity significantly moderated any connectivity-response associations within this network, using a threshold of *p_FDR_*<0.05. In addition, we calculated degree centrality for each network node (number of edges connected to it). We then examined correlations between degree and physiological response magnitude to identify nodes that are particularly central within the network as it relates to response magnitude. Degree was calculated on adjacency matrices thresholded at *r*=0.25 to avoid sparse connections. In all analyses, age was entered as a nuisance covariate.

## Results

### Threat-anticipatory physiological response and anxiety severity

Fig. 2A depicts mean anticipatory physiological response by cue, bin, and group (see Fig. S1 for individual-subject data). Analysis of physiological response during the 8s anticipation period indicated a significant Cue×Imminence interaction effect on SCL, *F*(3,144)=35.15, *p*<0.001, 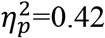, whereby the magnitude of SCL increased with imminence of the threat outcome, *F*(3,147)=31.48, *p*<0.001, 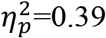, and decreased with imminence of the safe outcome, *F*(3,147)=6.33, *p*<0.001, 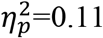. Across the sample, differential threat vs. safety responding significantly increased with outcome imminence, starting with absence of threat discrimination during initial response to the cues (bin 1), *t*(49)=0.60, *p*=0.55, *d*=0.06, and reaching maximal differentiation during *circa-strike* (bin 4), *t*(49)=5.70, *p*=6.7×10^−7^, *d*=0.88 (*t*-tests for dependent samples). Thus, the magnitude of anticipatory physiological response positively scaled, and showed greater threat specificity, with threat imminence.

**Figure 2.**
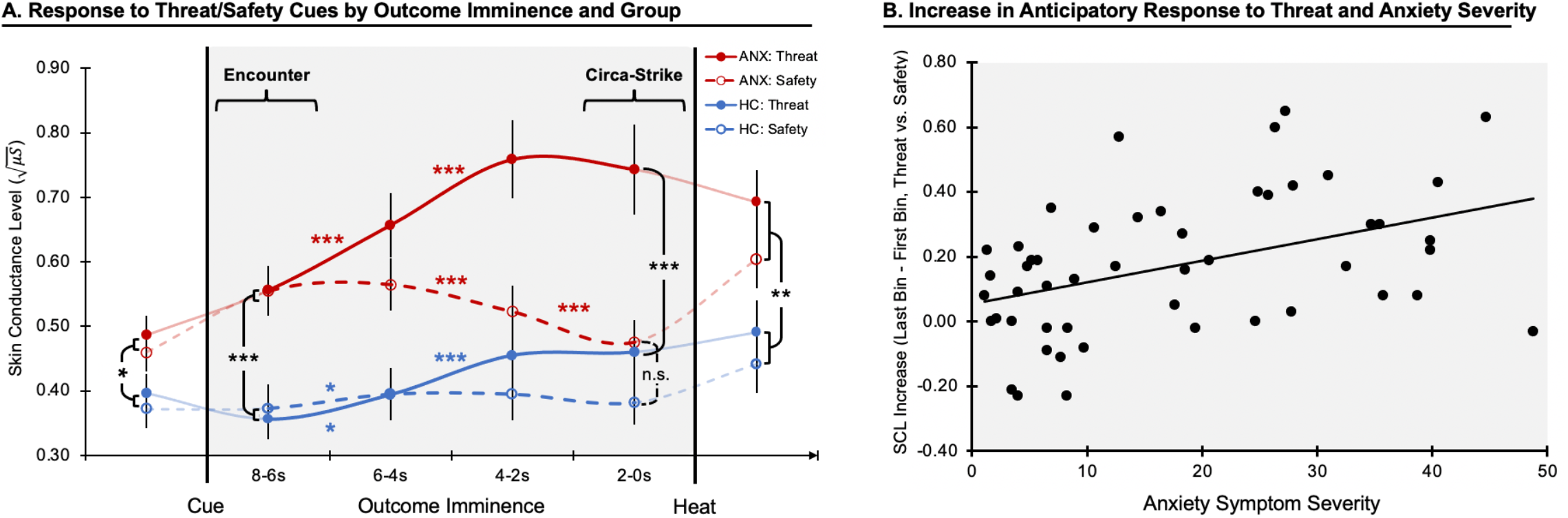
Changes in anticipatory physiological response by threat imminence. A) Area in gray represents anticipation period beginning with cue onset and up to imminent thermal stimulation. Lines reflect average skin conductance level in 2s bins in response to threat (highly painful heat, continuous lines) and safety (non-painful heat, dashed line), for the anxiety (red, ANX) and healthy control (blue, HC) groups. For completeness, we display responses prior to cue onset and during thermal stimulation. B) Scatterplot depicts association between anxiety symptom severity (assessed as a continuous measure) and magnitude of change in anticipatory physiological response to threat vs. safety (last bin minus first bin). *Note*: *, *p*<0.05, **, *p*<0.01, ***, *p*<0.001. Colored asterisks reflect within-group effects; black asterisks reflect between-group effects. Change in response over time is represented as curved lines for descriptive purposes, to reflect its assumed continuous nature.

This two-way interaction was qualified by a significant Cue×Imminence×Group interaction, *F*(3,144)=7.55, *p*<0.001, 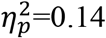, (Fig. 2A), indicating distinct temporal dynamics of physiological responding as a function of anxiety. This interaction effect was decomposed in ways that considered our hypotheses. First, we examined anxiety effects on *initial* responses to threat cues (*encounter* phase, first bin following cue onset). We noted a significant effect of Group, *F*(1,48)=17.05, *p*<0.001, 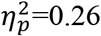, reflecting stronger initial responses across cues in the anxiety group. However, no Cue×Group effect was observed, *F*(1,48)=0.45, *p*=0.50, 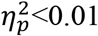, indicating that greater *encounter* response in anxiety is not specific to threat.

Second, we explicated anxiety differences in the unfolding of anticipatory response with threat imminence. In the anxiety group, a significant Cue×Imminence interaction was noted, *F*(3,72)=32.37, *p*<0.001, 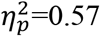, with follow-up analysis indicating that, relative to cue onset, response magnitude increased with threat imminence, *F*(3,72)=21.55, *p*<0.001, 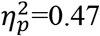, and decreased with imminence of the safe outcome, *F*(3,72)=13.39, *p*<0.001, 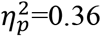. During *circa-strike*, when the outcome was most imminent, physiological responding in the anxiety group was significantly higher when threat vs. safety outcome was expected, *t*(24)=6.68, *p*=6.6×10^−7^, *d*=1.33. In the HC group, we noted weaker response discrimination over time, *F*(3,72)=6.22, *p*=0.001, 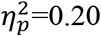, with response increasing in threat trials, *F*(3,72)=11.63, *p*<0.001, 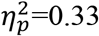, but not changing in safety trials, *F*(3,72)=0.82, *p*=0.49, 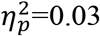, and a modest imminent threat vs. safety difference during *circa-strike, t*(24)=2.09, *p*=0.048, *d*=0.42. Comparing the groups during *circa-strike*, the anxiety group demonstrated a stronger response than the healthy group when anticipating immediate threat, *t*(48)=3.79, *p*<0.001, *d*=1.08, but not safety, *t*(48)=1.96, *p*=0.056, *d*=0.51. Finally, as per our hypothesis, we quantified the magnitude of *change* in threat-anticipatory response (last bin minus first bin in threat trials, relative to last bin minus first bin in safety trials). The anxiety group demonstrated a greater *threat-specific* increase in physiological response with imminence, *F*(1,48)=7.77, *p*=0.008, 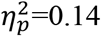. We assessed the internal reliability of this index; Cronbach’s alpha was 0.77, indicating acceptable to good reliability.

Thus, pathological anxiety was associated with a stronger initial response to cue onset which was not specific to threat. With increasing outcome imminence, anxiety was associated with greater anticipatory response magnitude to threat vs. safety.

To complement these group analyses, we examined whether the magnitude of initial response (bin 1) and evolving response (bin 4 minus bin 1) to anticipated threat was associated with individual differences in anxiety severity assessed continuously, in support of a dimensional mechanistic perturbation^84^. Anxiety symptom severity (as measured using total SCARED scores) was positively correlated with initial response to threat cues, *r*(48)=0.551, *p*<0.001, and safety cues, *r*(48)=0.437, *p*=0.001, but not to threat vs. safety difference (difference in bin 1 response magnitudes), *r*(48)=0.119, *p*=0.41. In terms of increasing anticipatory response, anxiety symptom severity was positively correlated with increase in response during anticipation of threat, *r*(48)=0.480, *p*<0.001, but not safety, *r*(48)=0.261, *p*=0.07, with the difference between these correlation coefficients being significant, *Z*=2.14, *p*=0.016. Finally, we quantified threat-specific increase in anticipatory response (bin 4 minus bin 1 in threat trials, relative to bin 4 minus bin 1 in safety trials). The correlation between anxiety severity and threat-specific increase in response was significant, *r*(48)=0.373, *p*=0.008; see Fig. 2B. These results therefore replicate the categorical analyses, indicating that anxiety along the severity continuum is associated with a threat-specific increase in anticipatory response with threat imminence. Results did not change after controlling for age. The threat-specific increase in response magnitude was then used in rsFC analyses (see below).

For completeness, we also report in the supplement on the Cue×Group effect during the period immediately before the cue-based anticipation window, and during thermal stimulation. Briefly, we observed a modestly greater pre-cue physiological response by the anxiety group (main effect); the primary interaction effect reported above (Cue×Imminence×Group) remained significant when controlling for pre-cue response. Response to thermal stimulation indicated greater response to the highly-painful heat relative to the non-painful heat, and a greater response by the anxiety group (main effect), although this effect was completely abolished once anticipatory response magnitude (bin 4) was controlled for. See supplement for full details.

### Intrinsic functional connectivity correlates of physiological response to anticipated threat

Imaging analyses focused on identifying network-level patterns of intrinsic functional connectivity that correlate with the magnitude of threat-anticipatory defensive responding. Of note, threat-specific anxiety effects emerged only when quantifying the magnitude of *increase* in physiological response with threat imminence across the anticipation window. As a result, we direct our analyses to identify rsFC correlates of this effect. To this end, we examined correlates of the threat-specific increase in anticipatory response, i.e., bin 4 minus bin 1 in threat trials relative to bin 4 minus bin 1 in safety trials, as defined above. rsFC associations with the magnitude of initial, non-threat-specific response are reported in Fig. S3.

Our primary analysis considered multivariate correlations among all edges in the defensive response network and physiological response magnitude, to identify subnetworks that most strongly relate to responding. This analysis revealed a specific subnetwork in which intrinsic connectivity significantly correlated with individual differences in response magnitude at the prespecified threshold, network *mass*=140.20, *p_FDR_*=0.035; see Fig. 3A (also see Fig. S2 for all edges at a lower significance threshold). The extent of connectivity within this subnetwork was associated with individual differences in physiological responding, as indicated by a significant correlation between its degree centrality and physiological response magnitude, *t*(33)=2.35, *p*=0.025 (Fig. 3B). This subnetwork comprised multiple positive cortical-subcortical connectivity-response edges, linking right vmPFC to bilateral BLA (*p*s<0.004) and hippocampus (*p*s<0.001), and to right NAcc (*p*=0.003), as well as positive left hippocampus-BNST connectivity (*p*=0.004). Connectivity values were predominantly positive in these edges. Among network nodes, right vmPFC demonstrated the strongest association between node degree and physiological response, *t*(33)=4.32, *p_FDR_*<0.001 (Fig. 3C).

**Figure 3.**
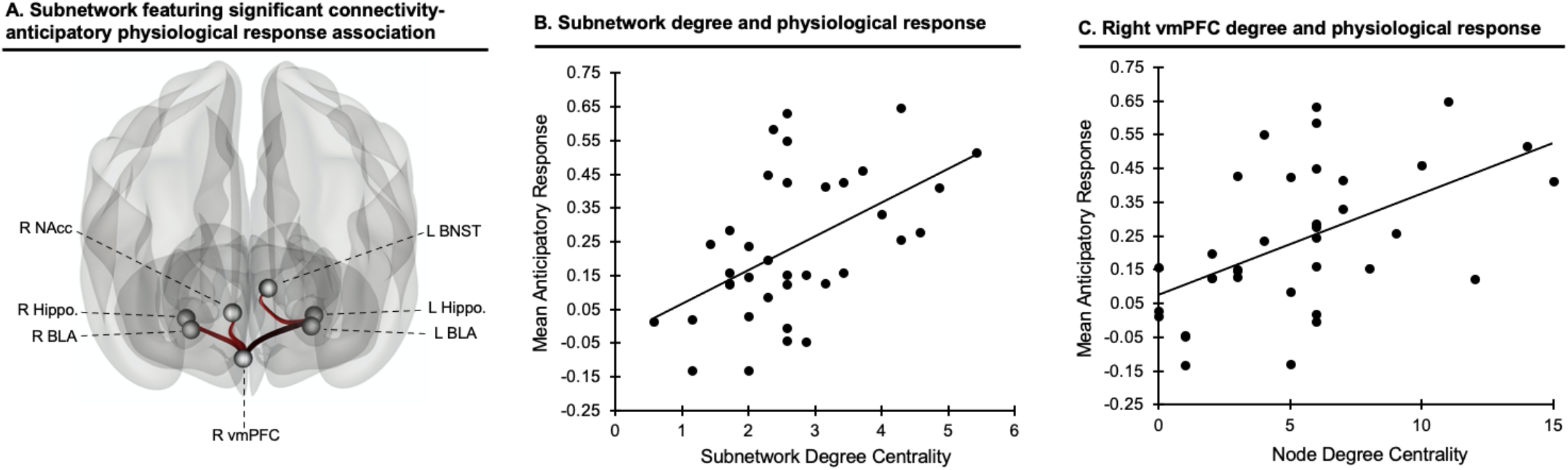
Subnetwork associated with threat-anticipatory physiological response. A) Functional connectivity within this subnetwork positively correlated with magnitude of anticipatory physiological response (*p_FDR_*<0.05). All depicted edges are positive. B) Significant association between the subnetwork’s degree centrality (average connectedness of nodes within it) and mean anticipatory physiological response magnitude; greater network connectedness was associated with greater response. C) Significant association between right vmPFC degree centrality and mean anticipatory physiological response magnitude; greater vmPFC connectedness to other nodes was associated with greater response. *Note*: R=right, L=left, NAcc=nucleus accumbens, BNST=bed nucleus of the stria terminalis, BLA=basolateral amygdala, hippo=hippocampus, vmPFC=ventromedial prefrontal cortex.

Finally, we examined moderation of connectivity-physiological response associations by anxiety severity in this subnetwork, using NBS and the same statistical threshold as above. A three-edge subnetwork, including right vmPFC links with bilateral hippocampus and left BLA, demonstrated a significant anxiety moderation effect of the association between connectivity and physiological response magnitude, *p_FDR_*=0.005, such that greater anxiety was associated with stronger connectivity-response correlation.

Additional analyses verified that the connectivity effects identified above did not reflect general covariation with non-specific physiological responsivity. Network analyses using the specified thresholds did not detect any networks in which connectivity was significantly associated with magnitude of physiological response during pre-cue (*pre-encounter*) or thermal stimulation, all *p_FDRs_*>0.05. Further, no significant associations emerged when threat imminence was not considered, i.e., when physiological response was averaged across bins in threat trials, or when the difference in averaged response in threat vs. safety trials was used in analysis, *p_FDRs_*>0.05. Thus, only when considering threat imminence did associations emerge between physiological response and function in the defensive response network.

For interested readers, we also report on auxiliary analyses detailing associations between function in this network and age and anxiety severity (using SCARED scores), regardless of physiological response (see Figures S4, S5). Edges were primarily negative in these analyses, indicating diminished connectivity with greater anxiety or age (although, notably, vmPFC edges were positive), although no significant subnetworks surpassed the specified significance threshold; see supplement.

## Discussion

This report incorporates insight from translational research on fear to characterize aberrant dynamics of threat-anticipatory physiological responding in anxiety. Analyses revealed three primary imminence-dependent findings. First, anxiety severity was positively associated with magnitude of physiological response to onset of both threat and safety cues. Second, anxiety moderated the temporal unfolding of anticipatory physiological response; with increasing imminence of physical threat, greater anxiety severity was associated with stronger anticipatory response. Finally, the magnitude of increase in threat-anticipatory response corresponded to intrinsic functional connectivity within a cortical-subcortical network, with vmPFC constituting a central node; more severe anxiety was associated with greater connectivity within this network as it relates to physiological response. Together, these findings link anxiety severity to unique temporal patterns of threat-anticipatory physiological response and variations in cortical-subcortical functional connectivity.

Anxiety symptoms are clinically characterized by excessive defensive responding in anticipation of threat^20^. Owing to the conserved nature of defensive responding, translational research on anxiety evokes threat-anticipatory states, primarily via threat learning or unpredictable threat paradigms. For example, studies of threat conditioning and physiology demonstrate anxiety-related enhanced skin conductance response to cues predicting potential threat, assessed over response windows of several seconds^24,85,86^, while other studies show potentiated eye-blink startle response in anxiety patients during more prolonged periods of anticipation of unpredictable threat^23,87,88^. Moreover, our recent work further localizes excessive physiological responding in anxiety to anticipation, but not ultimate experience, of aversive stimuli^53,89^. Such work establishes links between aberrant threat anticipation processes and anxiety symptoms, but does not consider threat imminence. Indeed, animal research suggests that an important determinant of defensive responding is proximity to threat. Research in healthy humans begins to extend this work, confirming that dynamic changes in physiological responding and defensive behaviors occur as a function of threat imminence. Given that threat-anticipatory defensive responding is central in the presentation of anxiety symptoms^20,21^, identifying these response dynamics could more precisely inform on psychopathological mechanisms. This is the first study to map dynamic patterns of imminence-induced changes in defensive responding to anxiety symptoms.

The primary dynamic effect that we observed was increasing physiological response as threat was becoming more imminent, in line with prior research^3,8,16,18,70^. In the context of research on defensive responding, this anticipation window corresponds to *post-encounter* to *circa-strike* phases. Threat imminence approaching *circa-strike* is associated with acute responding: physiological response discharge and increased execution of acute defensive behaviors, such as active avoidance/escape. Indeed, we observed an increase in physiological response as highly-painful, but not non-painful, thermal stimulation became more imminent. However, we extend such basic-science and translational research by demonstrating that anxiety moderates this effect. Specifically, relative to safety, greater evolving response to threat was observed in anxiety, culminating in a maximal anxiety effect during *circa-strike*, whereby individual with anxiety relative to healthy controls demonstrated greater response only when threat was most imminent. Thus, our findings highlight links between anxiety severity and perturbations in the mechanisms generating acute responding with increasing threat imminence.

Extensive research in animals identifies the brain circuitry underlying acute, threat-imminent defensive responding. Such research typically induces activation or deactivation in specific structures and measures changes in defensive behaviors, and indicates the prominent involvement of BLA, CeA, hippocampus, hypothalamus, and PAG, and their modulation by medial PFC^1,6,8,11,52,90^. Specifically, a circuit comprising vmPFC, BLA, and hippocampus has been identified as promoting the expression of excessive fear responses and anxiety-like behaviors in animals^28,91-95^. This distributed functional circuit is subserved by bidirectional functional and anatomical connectivity among these structures^29,95-98^. The involvement of these structures in the expression of defensive responses, including the expression of physiological responses, has recently been extended to humans^17-19,29,70,71,74,99-101^. Here, we further extend such work by using network analyses to characterize the multivariate nature of this circuit’s function as it corresponds to the expression of physiological response to threat. Importantly, we link perturbations in this circuit to anxiety severity. These links provide support for translational conceptualizations of anxiety implicating aberrant function of the vmPFC-BLA-hippocampus circuit in dysregulated fear responses observed in anxiety^8,93,95,102,103^. These effects emerged on resting-state connectivity patterns collected separately from the task, indicating a potential bias in this network’s intrinsic function that is clinically meaningful^104^.

Within this circuit, vmPFC centrality significantly related to physiological response magnitude. Considerable research in humans and animals highlights the role of vmPFC in threat processing and fear/anxiety states. For example, regulation of fear responses to extinguished threat has been shown to depend on vmPFC-BLA connections^102,105^, with the latter mediating downstream execution of acute defensive behaviors^90,106^. Likewise, vmPFC-ventral hippocampus connectivity has been implicated in the expression of fear responses^92,95,107^. More broadly, a recent conceptualization implicates vmPFC, and particularly its posterior extent, in threat (but not safety) assessment^108^. Our findings are in line with this conceptualization, as functional connectivity with this posterior region was highly correlated with the magnitude of expressed response to threat. The exact role that vmPFC plays in this context is still not clear, with some research suggesting that vmPFC exerts top-down response regulation in this circuit^29,102,109^, while others propose that vmPFC maintains fear representations potentially driven by bottom-up innervation^29,95,98^. While our findings cannot conclusively support a specific narrative, our observation of a positive association between connectivity-response correlation and anxiety severity, coupled by predominantly positive connectivity coefficients, are perhaps more in line with the latter notion^29^.

In addition, BNST-hippocampus connectivity was part of the subnetwork in which function correlated with physiological responding. BNST has been implicated as a key structure in defensive responding^9,22^, with studies demonstrating its function in response to both imminent and sustained threat^110-112^. Specifically, BNST-hippocampus connectivity has been suggested to modulate fear responding by integrating contextual information^113^. Here, increasing threat imminence may potentially reflect a contextual shift as the threat is increasingly temporally proximal, invoking this functional link. Additionally, we identified vmPFC-NAcc connectivity in the subnetwork. NAcc is ascribed a central role in mediating the behavioral aspects of defensive responses in face of imminent threat, and active avoidance behavior in particular^103,106,114^. Thus, enhanced NAcc connectivity may suggest an increased preparation to execute acute defensive behaviors with threat imminence. Interestingly, while research identifies CeA and PAG as outputs of this network in generation of defensive responses^6,70,94^, these structures did not significantly emerge in our data. This might mean that individual differences in defensive responding tendencies manifest primarily in variation in function of upstream network components in which inputs from multiple structures converge to influence the selection and maintenance of defensive responses, rather than in network effectors mediating their immediate execution^1^.

Contrary to our hypothesis, anxiety severity was not associated with greater threat-specific initial response to cue onset. In the context of potential encounter with threat, cue onset may correspond to the *encounter* phase (or start of the *post-encounter* phase)^8^. Our findings indicate that anxiety is associated with an elevated physiological response in this phase, but the initiation of this response precedes discrimination of threat vs. danger. Considering that physiological responding is meant to facilitate defensive behaviors, this enhanced response could reflect a drive to carry out freezing or passive avoidance behavior with cue onset, as an initial risk assessment process. Only following initial assessment of potential threat value, and as outcome becomes increasingly imminent, does physiological response show threat discrimination. An absence of a threat-specific effect on initial response in anxiety may contrast with theories emphasizing a bias towards threat in early, automatic attention allocation in anxiety^115^. Reconciling this apparent discrepancy, recent evidence suggests anxiety effects may manifest more reliably in later rather than earlier attention processes^116^. Considering threat imminence chronometry, cue onset occurs against the backdrop of a *pre-encounter* phase, in which subjects await the next trial. It is possible that in the context of *pre-encounter*, the onset of any stimulus that may predict danger initiates in individuals with anxiety a stronger risk assessment response. Indeed, in recent work, we showed that anxiety is associated with enhanced physiological response to both safety and threat cues^89^.

Extensive research in recent years attempts to identify pathophysiological markers for psychiatric disorders which could promote improved prediction, diagnosis, and treatment development^49,117,118^. The findings presented here, particularly the increase in physiological response to threat, provide initial indications for an anxiety biomarker, and encourage continued research for a number of reasons. First, threat-specific increase in anticipatory physiological response differentiated the healthy and anxiety groups. Second, increased physiological response provides face validity for this potential biomarker as acute physiological response to imminent is central in the presentation of anxiety symptoms^20,21^; further, this increase in response showed adequate internal consistency. Third, the association between response and anxiety severity also manifested dimensionally, in line with recent conceptualizations of psychopathology^84^; this could potentially offer greater sensitivity for identifying at-risk, sub-threshold individuals^118^.

Fourth, we linked this index to specific patterns of functional connectivity that correspond to findings from animal research; this could promote continued cross-species research on circuit-based biomarkers and treatment^118^. Finally, we observed these effects in youth with only anxiety disorders; this suggests specificity as well as potential utility for developmental samples. While these findings encourage additional research, additional work is required, including replication and continued assessment of reliability, validity, and specificity.

These findings should be considered in light of several limitations. First, sample size was modest; while effects were generally robust and emerged using continuous measures, a replication of these findings is nonetheless advised, especially for the imaging analyses. Second, we did not study youth with disorders other than anxiety; such samples are needed to establish the specificity of effects more conclusively. Third, imaging analyses relied on a 3T scanner; future work using higher fields could provide better image resolution and delineation of smaller subcortical structures. Finally, we used skin conductance as index of defensive responding; future research may wish to add additional physiological measures as well as assess continuous anxiety levels reports throughout the task and measure defensive behaviors^14^.

In conclusion, this report links translation research on fear, clinical research, and physiological and neuroimaging recording to identify aberrant patterns of threat imminence-dependent anticipatory responding in pathological anxiety. By considering threat imminence, we quantify a physiological index of anticipatory defensive responding that robustly differentiates anxious and healthy individuals, and has intrinsic functional connectivity correlates that correspond to findings from animal studies on defensive responding circuitry. These findings advance our understanding of normative and abnormal threat-anticipatory fear responses and establish a potential biomarker for anxiety.

## Data Availability

Limited data may be shared due to IRB rules regarding patient consent.

## Acknowledgements

We would like to thank Kalina J. Michalska, Elizabeth Necka, Chika Matsumoto, and Esther E. Palacios-Barrios for their assistance. We also thank the participants and families, as well as the staff of the Intramural Research Program of the National Institute of Mental Health (IRP, NIMH), National Institutes of Health. This research was supported (in part) by the NIMH IRP (ZIAMH002781-15, NCT00018057).

## Author contributions

1. Rany Abend: Contributed to conception and design, acquisition of data, analysis and interpretation of data; drafted the article and revised it critically for important intellectual content; gave final approval of the version to be published.
2. Sonia G. Ruiz: Contributed to acquisition of data, analysis and interpretation of data; drafted the article and revised it critically for important intellectual content; gave final approval of the version to be published.
3. Mira A. Bajaj: Contributed to acquisition of data, analysis and interpretation of data; drafted the article and revised it critically for important intellectual content; gave final approval of the version to be published.
4. Anita Harrewijn: Contributed to acquisition of data, analysis and interpretation of data; revised it critically for important intellectual content; gave final approval of the version to be published.
5. Julia O. Linke: Contributed to analysis and interpretation of data; revised it critically for important intellectual content; gave final approval of the version to be published.
6. Lauren Y. Atlas: Contributed to conception and design, interpretation of data; drafted the article and revised it critically for important intellectual content; gave final approval of the version to be published.
7. Daniel S. Pine: Contributed substantially to conception and design, analysis and interpretation of data; drafted the article and revised it critically for important intellectual content; gave final approval of the version to be published.

## Footnotes

1 Of note, this quantification measure correlated very highly, *r_s_*>0.90, *p_s_*<3.9×10^−19^, with alternative indices, such as difference between slopes of change and difference between raw responses in bin 4.

